# Predicting radiological severity of pulmonary tuberculosis in children: an assessment of the WHO-criteria and novel prediction scores on an individual participant dataset

**DOI:** 10.64898/2026.07.07.26357441

**Authors:** Akshita Gupta, Marieke M. van der Zalm, Minh Huyen Ton Nu Nguyet, Marc d’Elbée, Peter J. Dodd, Megan Palmer, Leyla Larsson, Alia Razid, Anneke C. Hesseling, Rory Dunbar, Norbert Heinrich, Heather J. Zar, Nyanda E. Ntinginya, Celso Khosa, Marriott Nliwasa, Valsan Philip Verghese, Maryline Bonnet, Eric Wobudeya, Bwendo Nduna, Raoul Moh, Juliet Mwanga-Amumpere, Ayeshatu Mustapha, Guillaume Breton, Jean-Voisin Taguebue, Laurance Borand, Pierre Goussard, H Simon Schaaf, Julie Morrison, Olivier Marcy, James A. Seddon, Chishala Chabala, Laura Olbrich, the Decide TB Study Group, the RaPaed-TB Consortium, the Umoya Study Consortium, and the TB Speed Consortium

**Affiliations:** Institute of Infectious Diseases and Tropical Medicine, LMU University Hospital, LMU Munich; German Centre for Infection Research (DZIF), Partner Site Munich, Munich, Germany; Desmond Tutu TB Centre, Department of Paediatrics and Child Health, Faculty of Medicine and Health Sciences, Stellenbosch University, Cape Town, South Africa; University of Bordeaux, National Institute for Health and Medical Research (Inserm) UMR 1219, Research Institute for Sustainable Development (IRD) EMR 271, Bordeaux, France; Sheffield Centre for Health and Related Research, School of Medicine & Population Health, University of Sheffield, Sheffield, United Kingdom; Clinical Research Department, Faculty of Infectious and Tropical Diseases, London School of Hygiene and Tropical Medicine, London, United Kingdom; Fraunhofer Institute ITMP, Immunology, Infection and Pandemic Research, Munich, Germany; Department of Paediatrics & Child Health, Red Cross War Memorial Children’s Hospital, MRC Unit on Child and Adolescent Health, University of Cape Town, South Africa; NIMR - Mbeya Medical Research Centre, P. O. Box 2410, Mbeya, Tanzania; Instituto Nacional de Saúde (INS), Maputo, Mozambique; Faculty of Medicine, Eduardo Mondlane University, Maputo, Mozambique; Kamuzu University of Health Sciences, Blantyre, Malawi; Pediatric Infectious Diseases, Christian Medical College, Vellore, India; TransVIMI, University of Montpellier, Institut de Recherche pour le Développement (IRD), Institut National de la Santé et Recherche Médicale (INSERM), Montpellier, France; MU-JHU Care Ltd, Kampala, Uganda; Arthur Davidson Children’s Hospital, Ndola, Zambia; Programme PAC-CI, Abidjan, Côte d’Ivoire; Epicentre Mbarara Research Centre, Mbarara, Uganda; Ola During Children Hospital, Fourah Bay Roadd, Freetown, Sierra Leone; Solthis, Paris, France; Mother and Child Center - Chantal Biya, Foundation, Yaoundé, Cameroon; Epidemiology and Public Health Unit, Institut Pasteur du Cambodge, Phnom Penh, Cambodia; Department of Paediatrics and Child Health, Stellenbosch University, Cape Town, South Africa; Department of Infectious Disease, Imperial College London, London, United Kingdom; University of Zambia, Department of Paediatrics and Child Health, School of Medicine, Lusaka, Zambia

**Keywords:** Treatment shortening, 4-month regimen, Treatment decision algorithm, paediatric, tuberculosis, pulmonary, treatment initiation, individual participant dataset, disease severity, prediction score, radiological severity

## Abstract

**Background:** The World Health Organization (WHO) recommends 4-month treatment for children with non-severe pulmonary tuberculosis, outlining eligibility criteria for settings with and without chest X-ray (CXR). We evaluated the diagnostic accuracy of the WHO eligibility criteria in settings without CXR (WHO-criteria) and developed clinical scores to support disease classification.

**Methods:** Using data from an individual participant dataset (IPD; Decide TB) of children with confirmed/unconfirmed tuberculosis from four diagnostic studies (RaPaed-TB, Umoya, TB-Speed HIV, TB-Speed Decentralisation), we assessed the diagnostic accuracy of the WHO-criteria (with/without bacteriological testing) using expert CXR interpretation as a reference. We developed two multivariable logistic regression models with (Score 1) and without (Score 2) bacteriological testing, converted coefficients into integer scores with a threshold of >10 corresponding to a sensitivity ≥70%.

**Results:** Of 2,383 children in the Decide TB IPD, 633 (26.6%) met the eligibility criteria for a 4-month regimen, of whom 116 (18.3%) had radiologically severe disease. With and without bacteriological testing, the WHO-criteria had sensitivities of 30.1% (95%CI: 20.3%-40.2%) and 21.7% (95%CI: 10.4%-34.5%), and specificities of 83.4% (95%CI: 80.2%-86.4%) and 81.9% (95%CI: 78.8%-84.9%), respectively. Score 1 and Score 2 had sensitivities of 41.1% (95%CI: 32.4%-49.5%) and 30.9% (95%CI: 22.6%-40.4%), and specificities of 77.3% (95%CI: 73.6%-80.8%) and 83.0% (95%CI: 79.5%-86.3%), respectively. Using WHO-criteria, 91/116 (78.4%) and 105/116 (90.5%) of children were at risk of undertreatment, compared to 68/116 (58.6%) and 80/116 (68.9%) when using developed scores.

**Conclusions:** Developed scores demonstrated better sensitivity than WHO-criteria, however, performance remained suboptimal. Implementing shorter antituberculosis regimens without CXR remains challenging in children.

## Introduction

The World Health Organization’s (WHO) Global Tuberculosis Report estimates that 1.2 million children and young adolescents (<15 years) developed tuberculosis in 2024, representing 11% of the global TB burden [1]. Two-thirds of children with pulmonary tuberculosis are estimated to have non-severe disease [2], most with smear-negative and non-severe forms of tuberculosis that could likely be treated for shorter durations [3]. Such treatment shortening may offer several advantages including reduced drug-induced adverse effects, improved adherence to treatment, reduction of costs of treatment to the family and the healthcare system [4–6].

In 2022, the SHINE trial demonstrated that a 4-month regimen of antituberculosis treatment was noninferior to a 6-month regimen in children with non-severe disease [3]. Based on these findings, the WHO issued operational guidelines outlining eligibility criteria for implementation of the 4-month regimen [4]. Classification of non-severe pulmonary tuberculosis under this guidance requires chest X-ray (CXR) evaluation to exclude radiologically severe disease [4]. Recognising that access to CXR may not be possible in many settings, the WHO provided additional recommendations for implementing the 4-month regimen where radiology was unavailable, (referred to as ‘WHO-criteria’) both with and without access to bacteriological testing (molecular testing and/or microscopy) (**Supplemental table 1**) [3,4]. In the absence of CXR, WHO recommends that children diagnosed with tuberculosis who present with mild symptoms not requiring hospitalization, with low bacillary burden (defined as smear negative or with semi-quantitative molecular testing results of negative, trace, very low or low) may be considered for the 4-month treatment regimen, provided these children are followed up monthly and respond well to treatment [4]. However, this recommendation is based on expert consensus with no evidence whether this approach ensures safe and effective treatment. This uncertainty poses challenges for implementation within national tuberculosis control programmes [6]. To facilitate implementation in low-resource settings, a data-driven approach predicting radiological severity of disease using available clinical indicators could complement existing WHO treatment decision algorithms (TDAs) in identifying children eligible for the 4-month regimen [4].

Using individual participant data (IPD) from children recruited to four existing well-characterised paediatric tuberculosis cohorts across eight countries [7], we sought to assess the WHO-criteria for starting a 4-month regimen with and without bacteriological testing. We developed two scores, with (Score 1) and without (Score 2) bacteriological testing to predict radiological severity of pulmonary tuberculosis in children.

### Methodology

#### Study Design, population, and establishment of an IPD

A large individual participant dataset (IPD) was developed within the Decide TB project (EDCTP101103283) that included data from 2,383 children from four diagnostic studies (RaPaed-TB [8], Umoya [9], TB-Speed HIV [10,11], and TB-Speed Decentralisation [12]). In short, the IPD was prepared by extracting variables included in the TDAs (e.g. presence of symptoms, tuberculosis exposure, HIV status) and conducting subsequent standardisation before merging into a single dataset, to ensure that standard scales and definitions were used, including the creation of composite variables. Further details of the project [13], individual studies and variables in the IPD have been published [7] and are summarised in **Supplemental table 2-3**.

The WHO eligibility criteria are guidance for initiating the 4-month regimen in children with pulmonary tuberculosis as outlined in Box 5.3 from Module 5 of the Operational Handbook on Tuberculosis [4]. For this study, the WHO eligibility criteria in settings without CXR facilities is referred to as ‘WHO-criteria’ (i.e., described as Setting B and Setting C in Box 5.3 from Module 5 of the Operational Handbook; Supplemental table 1) [4]. Based on the WHO-criteria, we included children between 3 months and 10 years of age who were retrospectively classified as having either confirmed, unconfirmed or unlikely tuberculosis using National Institute of Health (NIH) 2015 clinical case definitions (**Supplemental table 4**) [14], and were eligible for the 4-month regimen based on clinical presentation as outlined in the WHO-criteria (i.e., non-severe tuberculosis, without features precluding 4-month regimen use). Eligibility for the 4-month regimen under WHO-criteria is not synonymous with non-severe disease; only a subset of children with non-severe tuberculosis qualifies, as those with advanced HIV or severe acute malnutrition (SAM) remain ineligible irrespective of disease severity.

Thus, we excluded children with unlikely tuberculosis, extrapulmonary disease manifestations, without available Xpert MTB/RIF or Xpert MTB/RIF Ultra results, and children with severe clinical presentation as defined in the WHO operational handbook, being children living with HIV (CLHIV) with CD4 counts <100 cells/mm3 or unknown HIV status, SAM, respiratory distress, high fever (≥39°C), severe pallor, restlessness, irritability or lethargy.

#### Radiographic Evaluations

As part of the original studies, all children had a CXR performed at baseline [7,15–17]. CXRs were retrospectively reviewed centrally by a single independent paediatric tuberculosis expert, to ensure uniformity across included studies. ‘Radiologically non-severe disease’ was defined using WHO definitions described in Setting A from in Box 5.3 from Module 5 of the Operational Handbook [4] as a) normal; or b) abnormal CXRs with any of the following features: intrathoracic lymph node tuberculosis without significant airway obstruction, or pulmonary tuberculosis confined to one lobe with no cavities and no miliary pattern, or uncomplicated pleural effusion (without pneumothorax or empyema). Abnormal CXRs with findings additional to those outlined (including multilobe presentations, airway obstruction, cavitary or miliary presentations, pneumothorax or empyema) were considered as ‘Radiologically severe disease’.

#### Data analysis and reference standards

All data management and analysis were conducted using R version 4.4.2/ RStudio version 2024.12.1. Socio-demographic characteristics were summarised using proportions for categorical variables, and medians with interquartile ranges (IQR) or means with standard deviations (SD) for continuous variables. To assess the sensitivity and specificity of the WHO-criteria, we used CXR readouts from a single blinded expert as the reference standard for diagnostic accuracy estimation based on the definitions outlined above. Sensitivities and specificities were reported with 95% confidence intervals (CI) based on bootstrapping 1000 times. This analysis was repeated for predefined subgroups of interest (parent study, age, NIH 2015 case definition, Weight/BMI-for-age Z scores [WAZ], HIV status, healthcare level [Primary versus District versus Tertiary level], history of TB exposure).

#### Prediction score development

We developed two multivariable logistic regression models with a random intercept to account for heterogeneity across parent studies: Model 1, including bacteriological testing, and Model 2, without bacteriological testing. The models included predictors from clinical features commonly considered during the evaluation of presumptive childhood pulmonary tuberculosis in primary healthcare settings and included in the WHO TDAs [4]. Details on the clinical predictors can be found in Supplemental table 3.

We performed internal validation for each model using bootstrap resampling 1000 times. The optimism estimate corresponded to the mean apparent-minus-test AUROC (area under the receiver-operating characteristic curve) difference across the bootstrap resamples. The optimism-corrected AUROC was calculated and reported as the apparent AUROC of the original fitted model minus the estimated optimism. We examined the ROC curves of both models to assess their discriminative ability. To facilitate the use of the model by clinicians, we translated the model into a scoring system, Score 1 (=Model 1) and Score 2 (=Model 2), where each predictor is assigned points on a scale of 10. We identified a range of different candidate logit thresholds (of predicted probabilities) corresponding to a sensitivity for classifying radiologically severe disease of at least 50%, and up to 90%. For each candidate threshold, we derived a scaling factor by subtracting the model intercept from the threshold and dividing by 10. Multiplying each predictor’s β coefficient by this scaling factor yielded its integer point value (see **Supplemental tables 5 and 6** for the final Score 1 and 2 development) [18]. We chose the final Score 1 and 2 after discussion based on: the balance between sensitivity and specificity for the threshold, the resulting integer values of each predictor after translation on a scale of 10, and final scores being practical for clinical use (see **Supplemental methods**).

Diagnostic accuracy was estimated for Score 1 and Score 2 compared to the reference standard as defined above, and mean sensitivities and specificities estimated through bootstrapping 1000 times were reported. Children at risk of undertreatment (false negatives) were defined as those children with radiologically severe disease who were incorrectly predicted as eligible for the 4-month regimen. Children at risk of overtreatment (false positives) were defined as those children with radiologically non-severe disease who were incorrectly predicted as eligible for the standard 6-month regimen.

## Results

### General population description

Of 2,383 children included in the original studies (RaPaed-TB: n=975, Umoya: n=547, TB-Speed HIV: n=277, TB-Speed Decentralisation: n=584), 792 (33.2%) were excluded due to missing data or lack of diagnostic case classification, and a further 958 (40.2%) were excluded because their clinical presentation did not meet WHO-criteria for the 4-month regimen (**Figure 1**). Among the 633 children included in this analysis, the median age was 2.8 years (IQR: 1.3 - 5.2), 294 (46.4%) were female, and 118 (18.6%) were CLHIV. In total, 202 (31.9%) had confirmed tuberculosis and 431 (68.1%) unconfirmed tuberculosis. Most children had radiologically non-severe disease (517; 81.7%) and low bacterial burden (598; 94.5%) with semiquantitative Xpert MTB/RIF or Ultra results as negative or below medium (**Table 1, Supplemental table 7**).

**Figure 1.**
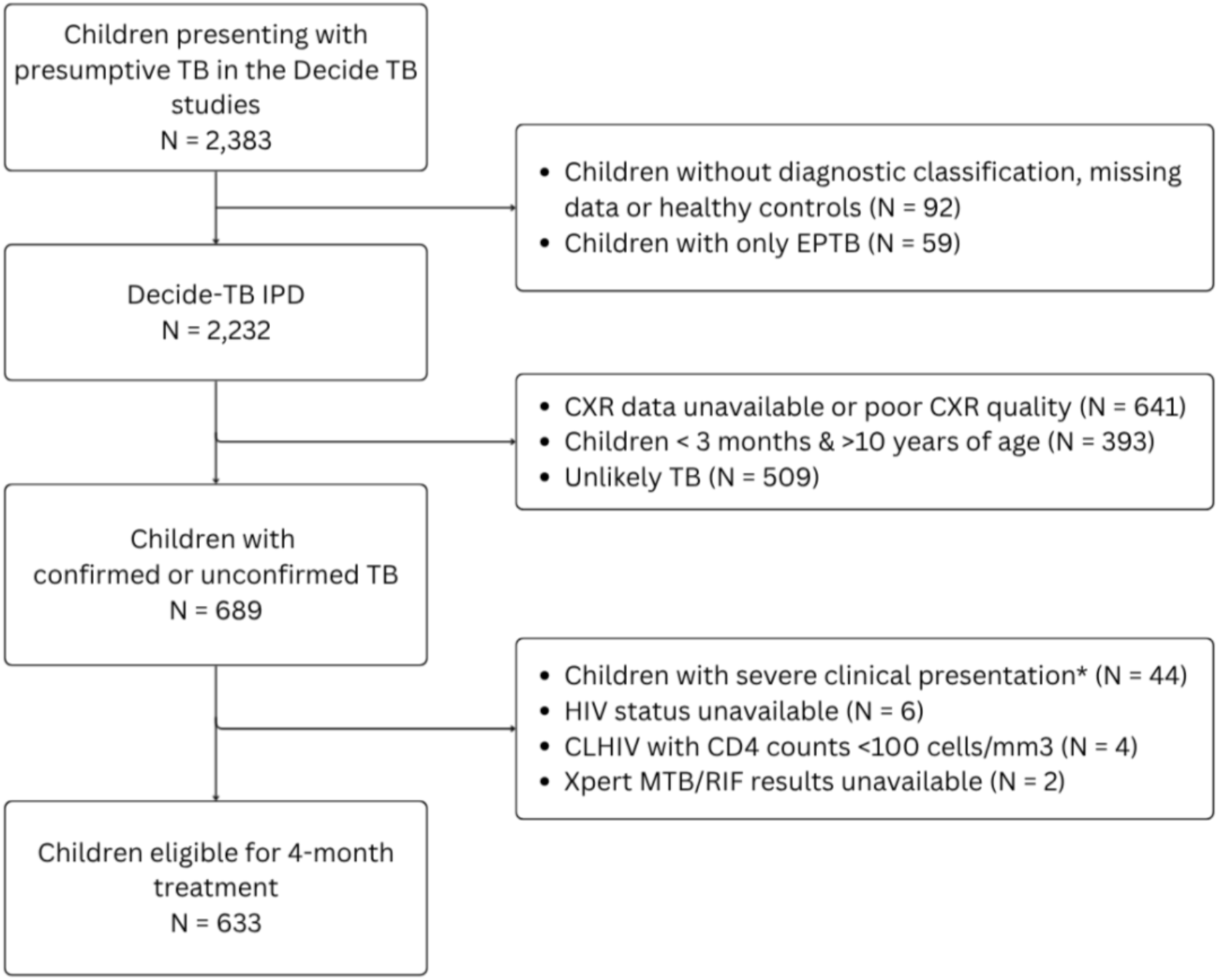
Development of the Decide TB individual participant dataset (IPD) for radiological severity prediction. Children were recruited from RaPaed-TB, Umoya, TB-Speed HIV, and TB-Speed Decentralization parent studies to build the Decide TB IPD. After applying inclusion and exclusion criteria, children who could be evaluated for 4-month regimen per the WHO eligibility criteria were included in the final analysis. *Eligibility for the 4-month regimen under the WHO criteria is not synonymous with non-severe disease as children may have non-severe TB and yet remain ineligible due to additional factors such as advanced HIV or severe acute malnutrition (SAM). Thus, we excluded children with severe clinical presentation as defined in the WHO operational handbook – severe acute malnutrition, respiratory distress, high fever (over 39°C), severe pallor, restlessness, irritability or lethargy [1]. IPD: Individual participant dataset; CXR: Chest X-ray; TB: Tuberculosis CLHIV: Children living with HIV. EPTB: extrapulmonary TB

**Table 1.** Characteristics of children included in the individual participant dataset by radiological disease severity#.

| Characteristic | All Children in this<br>IPD<br>N = 633 | Radiologically non-severe disease<br>N = 517 (81.7%) | Radiologically severe<br>disease<br>N = 116 (18.3%) |
| --- | --- | --- | --- |
| **Study** |  |  |  |
| RaPaed-TB (%) | 272 (43.0%) | 198 (38.3%) | 74 (63.8%) |
| Umoya (%) | 163 (25.8%) | 146 (28.2%) | 17 (14.7%) |
| TB-Speed HIV (%) | 55 (8.7%) | 47 (9.1%) | 8 (6.9%) |
| TB-Speed Decentralization (%) | 143 (22.6%) | 126 (24.4%) | 17 (14.7%) |
| **Age (years) [median (IQR)]** | 2.81 (1.26, 5.21) | 2.82 (1.35, 5.29) | 2.67 (0.97, 5.07) |
| **Age (categories)** |  |  |  |
| 3 months to <2 years (%) | 249 (39.3%) | 198 (38.3%) | 51 (44.0%) |
| 2 to <5 years (%) | 214 (33.8%) | 178 (34.4%) | 36 (31.0%) |
| 5 to <10 years (%) | 170 (26.9%) | 141 (27.3%) | 29 (25.0%) |
| **Sex** |  |  |  |
| Female (%) | 294 (46.4%) | 239 (46.2%) | 55 (47.4%) |
| Male (%) | 339 (53.6%) | 278 (53.8%) | 61 (52.6%) |
| **Weight/BMI for Age (Z score)** |  |  |  |
| > -1 (%) | 341 (53.9%) | 283 (54.7%) | 58 (50.0%) |
| > -2 to < -1 (%) | 187 (29.5%) | 153 (29.6%) | 34 (29.3%) |
| > -3 to < -2 (%) | 105 (16.6%) | 81 (15.7%) | 24 (20.7%) |
| **CLHIV (%)** | 118 (18.6%) | 91 (17.6%) | 27 (23.3%) |
| **NIH 2015 case definition** |  |  |  |
| Confirmed tuberculosis (%) | 202 (31.9%) | 149 (28.8%) | 53 (45.7%) |
| Unconfirmed tuberculosis (%) | 431 (68.1%) | 368 (71.2%) | 63 (54.3%) |
| **Facility type** |  |  |  |
| Primary Healthcare (%) | 4 (0.6%) | 4 (0.8%) | 0 (0.0%) |
| District Healthcare (%) | 139 (22%) | 122 (23.6%) | 17 (14.7%) |
| Tertiary Healthcare (%) | 490 (77.4%) | 391 (75.6%) | 99 (85.3%) |
| **Symptoms** |  |  |  |
| Cough ≥ 2 weeks (%) | 427 (67.5%) | 351 (67.9%) | 76 (65.5%) |
| Fever ≥ 2 weeks (%) | 256 (40.4%) | 211 (40.8%) | 45 (38.8%) |
| Weight loss (%) | 358 (56.6%) | 282 (54.5%) | 76 (65.5%) |
| Lethargy (%) | 276 (43.6%) | 206 (39.8%) | 70 (60.3%) |
| Haemoptysis (%) | 14 (2.2%) | 7 (1.4%) | 7 (6.0%) |
| Night sweats (%) | 220 (34.8%) | 185 (35.8%) | 35 (30.2%) |
| Enlarged lymph nodes (%) | 110 (17.4%) | 85 (16.4%) | 25 (21.6%) |
| Tachycardia (%) | 53 (8.4%) | 35 (6.8%) | 18 (15.5%) |
| Tachypnoea (%) | 103 (16.3%) | 74 (14.3%) | 29 (25.0%) |
| **TB-related findings** |  |  |  |
| TB Exposure (%) | 360 (56.9%) | 306 (59.2%) | 54 (46.6%) |
| Anti-tuberculosis treatment initiated (%) | 592 (93.5%) | 482 (93.2%) | 110 (94.8%) |
| **Bacteriological Testing** |  |  |  |
| **Xpert MTB/RIF or Ultra** |  |  |  |
| *Mtb* Negative (%) | 494 (78.0%) | 417 (80.7%) | 77 (66.4%) |
| *Mtb* Positive, including trace (%) | 139 (22.0%) | 100 (19.3%) | 39 (33.6%) |
| **Ziehl-Neelsen smear** |  |  |  |
| Negative (%) | 217 (94.3%) | 187 (96.9%) | 30 (81.1%) |
| Positive (%) | 13 (5.7%) | 6 (3.1%) | 7 (18.9%) |
| Not performed (%) | 403 | 324 | 79 |
| **WHO eligibility criteria groups** |  |  |  |
| Xpert MTB/RIF or Ultra results:<br>Negative, Trace, Low, very low<br>OR<br>smear negative (%) | 598 (94.5%) | 499 (96.5%) | 99 (85.3%) |
| Xpert MTB/RIF or Ultra results:<br>Medium, high, very high<br>OR<br>smear positive (%) | 35 (5.5%) | 18 (3.5%) | 17 (14.7%) |

### Evaluation of the WHO-criteria for classifying radiologically severe disease

The WHO-criteria with bacteriological testing had a sensitivity of 30.1% (95%CI: 20.3%-40.2%) and a specificity of 83.4% (95%CI: 80.2%-86.4%) for classifying radiologically severe disease. The WHO-criteria without bacteriological testing had a sensitivity of 21.7% (95%CI: 10.4%-34.6%) and a specificity of 81.9% (95%CI:78.8% - 84.9%). The WHO-criteria showed consistently low sensitivities across all subgroups, with and without bacteriological testing, with the lowest values observed in children aged 2 to 5 years (10.9% and 5.4% respectively) and those with WAZ > −2 to < −1 (14.4% and 2.8%). Irrespective of bacteriological testing, children without a history of TB exposure had higher sensitivity for radiologically severe disease than those with TB exposure (with bacteriology sensitivity of 27.3% vs 14.8% in tuberculosis-exposed subgroup; without bacteriology sensitivity of 16.0% vs 1.8% in tuberculosis-exposed subgroup; **Supplemental Tables 8-9**).

### Development and evaluation of prediction scores

Model 1 was developed for settings with bacteriological testing and Model 2 for those without bacteriological testing available. Model 1 had an apparent AUROC of 0.732, with a mean optimism of 0.0234 (95%CI: −0.027 to 0.071), yielding an optimism-corrected AUROC of 0.708. Model 2 had an apparent AUROC of 0.699, with a mean optimism of 0.0226 (95%CI: −0.021 to 0.071), yielding an optimism-corrected AUROC of 0.676. We developed Score 1 based on Model 1 and Score 2 based on Model 2 (**Figure 2**; Supplemental Tables 5 and 6). The cut-off selection was driven by the balance between sensitivity and specificity for the threshold, the resulting integer values of each predictor after translation on a scale of 10, and final scores being practical for clinical use. For Score 1, we selected a cut-off on the ROC curve of Model 1 corresponding to a sensitivity of 75.9% (95%CI: 67.3-82.7) and a specificity of 61.5% (95%CI: 57.2-65.6); for Score 2, a cut-off on the ROC curve of Model 2 corresponding to a sensitivity of 68.9% (95%CI: 60.1-76.7) and a specificity of 64.8% (95%CI: 60.6-68.8) for the detection of radiologically severe pulmonary tuberculosis (Figure 2).

**Figure 2.**
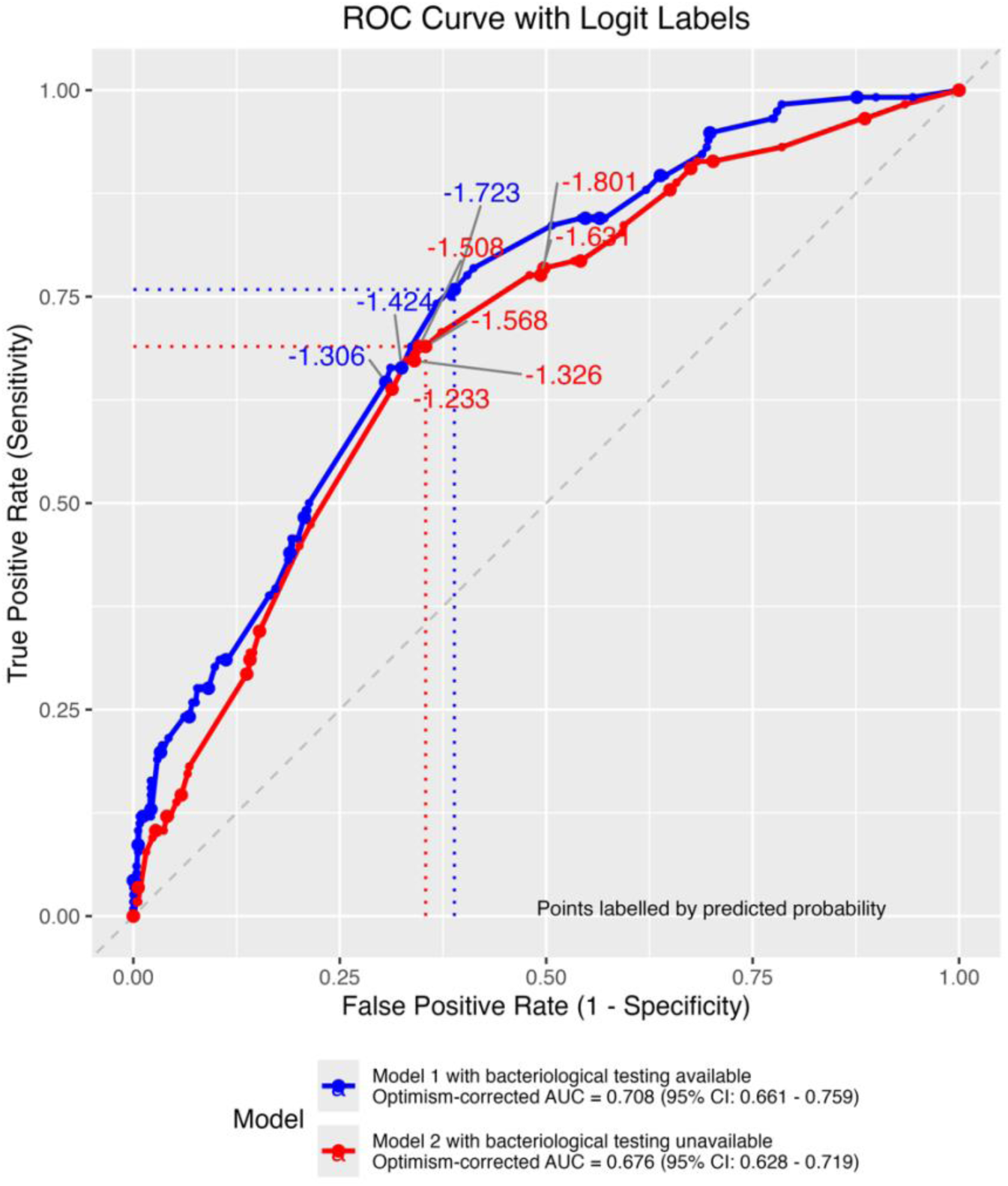
Receiver operating characteristic (ROC) curve for model 1 and model 2 in settings for logit selection and score development in dataset. (1) model 1 (blue) integrated all predictors including quantitative Xpert MTB/RIF or Ultra results. Optimism corrected AUC of model 1 was 0.708 (95%CI: 0.66-0.76), (2) Model 2 (red) excluded quantitative Xpert MTB/RIF or Ultra results. Optimism corrected AUC of model 2 was 0.676 (95%CI: 0.63-0.72). Points on the ROC curve indicate the candidate logit thresholds (predicted probabilities) of the model. A sensitivity of >70% was taken for selecting the cut off for the developed scores: −1.723 for model 1 (blue dotted line) and −1.568 for model 2 (red dotted line). AUC: Area under the curve

On diagnostic accuracy evaluation using the development dataset, Score 1 had a mean sensitivity of 41.1% (95%CI: 32.4%-49.5%) and mean specificity of 77.3% (95%CI: 73.6%-80.8%); while Score 2 had a mean sensitivity of 30.9% (95%CI: 22.6%-40.4%) and a mean specificity of 83.0% (95%CI: 79.5%-86.3%). The scores were evaluated among different subgroups of interest (**Supplemental Tables 10-11**). Across both scores, WAZ >−3 to <−2 was consistently associated with higher sensitivity (Score 1, 50.6% and Score 2, 42.3%) but the lowest specificity of any subgroup (Score 1, 65.2% and Score 2, 72.2%), and children without a history of tuberculosis exposure had higher sensitivity than those with known exposure in both scores (Score 1 sensitivity of 53.3% vs 28.0% in tuberculosis-exposed subgroup; Score 2 sensitivity of 40.6% vs 20.5% in tuberculosis-exposed subgroup). A schematic representation of how these scores could be incorporated into the WHO TDAs is shown in **Supplemental Figure 1** and displays the score assigned to each clinical variable included in the models.

### Over- and undertreatment based on predicted radiological disease severity

The WHO-criteria with bacteriological testing correctly identified 460/517 (88.9%) children with non-severe and 25/116 (21.6%) children with severe disease. Score 1 correctly identified 400/517 (77.4%) children with non-severe disease and 48/116 (41.4%) children as severe disease (**Figure 3A**, Supplemental Tables 8 and 10). Among children with severe disease, the WHO-criteria incorrectly identified 91/116 (78.4%) children as non-severe and Score 1 incorrectly identified 68/116 (58.6%) children as non-severe, placing them at risk of undertreatment.

**Figure 3.**
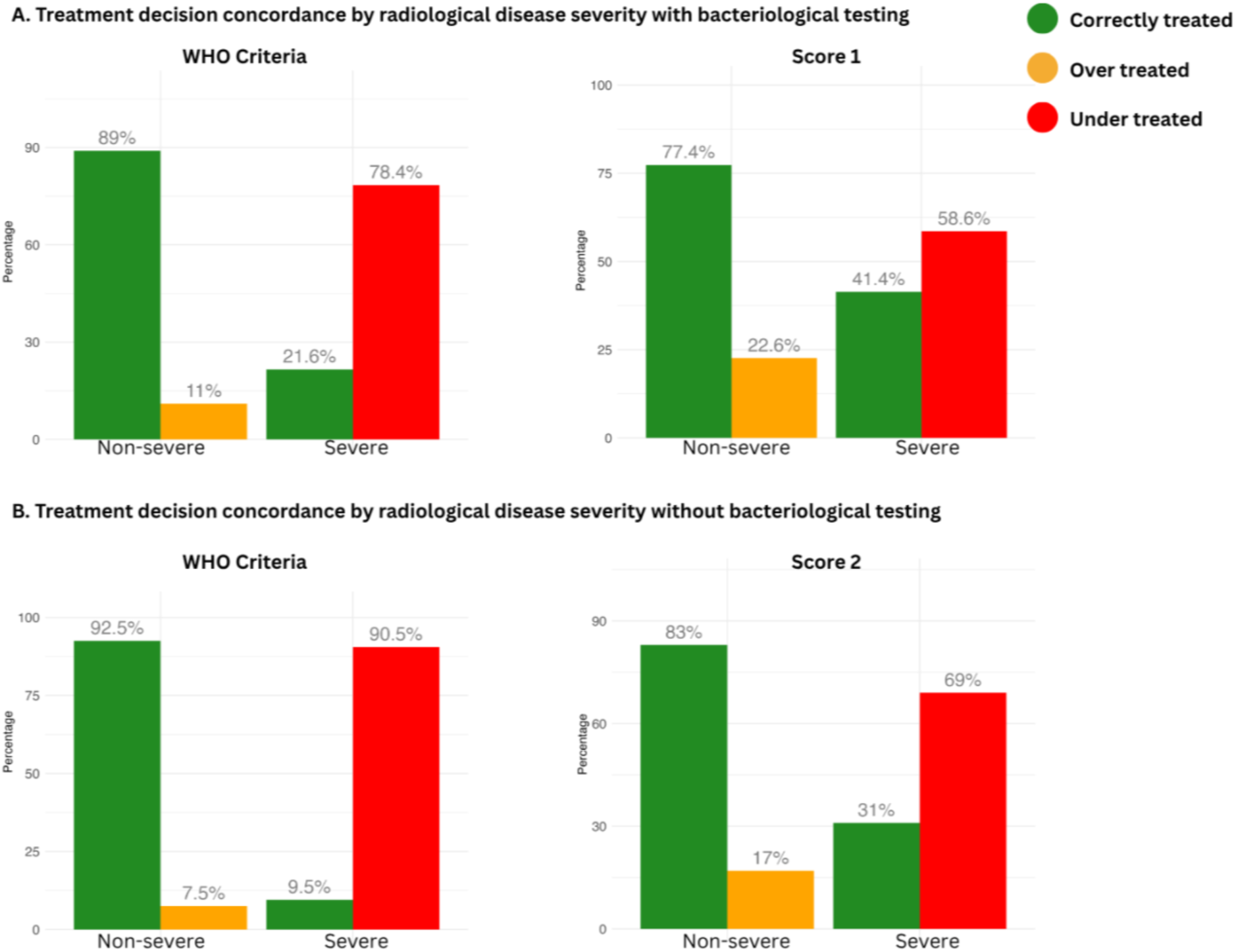
Treatment decision concordance in children predicted as severe or non-severe based on the WHO criteria and developed scores considering with and without bacteriological testing. Where Y-axis is the percentage of children and x-axis is the number of true radiologically non-severe (n = 517) and severe chest X-ray features (n = 116). (A) Classification of children based on the WHO-criteria and developed Score 1 with bacteriological testing (Setting B WHO Eligibility Criteria). (B) Classification of children based on the WHO-criteria and developed Score 2 without bacteriological testing (Setting C WHO Eligibility Criteria).

The WHO-criteria without bacteriological testing correctly identified 478/517 (92.5%) children with non-severe and 11/116 (9.5%) of children with severe tuberculosis disease. Score 2 correctly identified 429/517 (82.9%) children as non-severe and 36/116 (31.0%) children as severe tuberculosis disease (**Figure 3B**, Supplemental Tables 9 and 11). The WHO-criteria led to 105/116 (90.5%) children incorrectly classified with severe disease, placing them at risk of undertreatment; while Score 2 led to 80/116 (68.9%) children incorrectly classified with severe disease, placing them at risk of undertreatment.

## Discussion

This is the first study to assess the performance of WHO eligibility criteria for the 4-month treatment regimen for settings where CXR is unavailable and to develop clinical prediction scores corresponding to radiological severity of pulmonary tuberculosis in children. The analysis leverages a large, well-characterised, and comprehensive IPD, covering a wide range of geographical regions with a high burden of tuberculosis. Our scores are intended for use alongside the WHO TDAs and to support identification of children eligible for the 4-month regimen when CXRs are not routinely available. While both scores improve radiological severity prediction compared with the current WHO-criteria, their overall performance remains suboptimal, resulting in a substantial risk of undertreatment among children with radiologically severe tuberculosis.

The WHO-criteria and Score 1 demonstrate better disease classification with bacteriological testing than without. However, given the paucibacillary nature of pulmonary tuberculosis in children [2,3] and the challenges in obtaining reliable respiratory samples [19], bacteriological confirmation is often not feasible. In such circumstances, disease severity would be evaluated primarily based on clinical symptoms, particularly in high-burden settings at lower (primary) levels of care [20]. The limitations of symptom-based severity classification are well-described, especially among children under 5 years in whom constitutional symptoms such as cough, fever, and weight loss are non-specific; overlapping with other common childhood illnesses and do not reliably reflect radiological tuberculosis disease severity [14,21–24].

Our analysis showed that using WHO-criteria or our scores carry a risk of misclassification leading to potential undertreatment of children with radiologically severe disease. The WHO-criteria address this concern by recommending a monthly follow up and allows for treatment to be extended to 6 months, if clinically indicated [4]. In the IPD overall, most children had unconfirmed tuberculosis and would require close follow-up during treatment. Furthermore, less than a third of children across the different studies had severe disease, in line with literature [2,3,5]. Taken together, these findings suggest that, although in practice the number of children at risk for undertreatment would be small, careful follow-up for response to therapy remains essential.

Based on our results, better access to affordable and interpretable radiology remains a critical need for the safe implementation of the 4-month regimen. In settings where radiological infrastructure exists but experienced CXR reviewers are lacking, computer-aided detection tools could potentially support severity assessment [25,26]. Such approaches could ultimately be used either to directly classify disease severity or to complement existing clinical decision-making tools such as the WHO TDAs or our scores [27]. Definitions of severe pulmonary tuberculosis in children rely largely on CXR features, and possible alternative approaches to disease classification methods that are less dependent on CXR may aid broader implementation of shorter regimens. Another consideration is the validity of radiological disease severity itself as a reference standard, as the SHINE trial enrolled children with only smear-negative, clinical and radiological non-severe disease. There is, however, no clear scientific evidence that the severe radiological features are indeed associated with more severe disease requiring longer TB treatment. Therefore, highlighting that there is no direct trial evidence that children with severe radiological features would fare worse on a shortened regimen. Taken together, these gaps point to a need for further research both on alternative approaches to severity classification and on the safety of shorter regimens across the full spectrum of disease severity.

Key limitations of this analysis are that the performance of our scores across subgroups was evaluated in the same dataset and external validation of the models in independent datasets was not performed. We used the retrospective NIH 2015 clinical case definitions for confirmed and unconfirmed tuberculosis as our inclusion criteria, as they provided a standardised, validated framework for tuberculosis classification enabling consistent groups across studies. Including all treated children would have introduced a heterogeneous population including potentially unlikely tuberculosis cases, risking contamination of the reference standard and biasing model performance. CXR interpretation forms part of the clinical case definitions which may introduce circularity when evaluating CXR-based predictors and potentially overstating utility. It is possible that the radiological features seen in some of the children classified as unconfirmed tuberculosis may have been attributable to alternative diagnoses, potentially impacting the accuracy of the models. In addition, the recruitment settings of the parent studies may limit generalisability, as most children were enrolled from district and tertiary level hospitals. This could potentially underrepresent primary level settings where a substantial burden of childhood tuberculosis exists, and radiological facilities or expert radiologists are often lacking. The CXR interpretation included in this analysis was performed by an experienced paediatric tuberculosis expert, which may not reflect the interpretation in routine clinical practice, where availability of experienced readers cannot always be assured. Lastly, this retrospective analysis could not assess the status of the child at the 4-month timepoint (reflecting end of treatment for non-severe disease) against the 6-month timepoint after starting treatment. Prospective studies comparing 4-month regimen outcomes are required to support implementation by national tuberculosis programs. Despite these limitations, our findings fill current knowledge gaps in implementing the 4-month regimen in children with pulmonary tuberculosis.

In conclusion, although our scores had better sensitivity than the current WHO-criteria, they are unable to reliably identify children with radiologically severe tuberculosis disease based on clinical features alone, with or without bacteriology. Implementing the 4-month regimen for children with pulmonary tuberculosis in the absence of adequate CXR infrastructure remains challenging and underscores the need for improved access to affordable and feasible CXR in resource-limited settings.

## Data Availability

The IPD will be made available upon reasonable request with investigator support and a signed data access agreement. Submit a request to Alberto Beyersdorff, data manager at LMU Munich. The code for the analysis is available at https://github.com/4Gupt4/Decide-TB.

## Acknowledgements

We would like to acknowledge all children and caregivers that made the original studies possible, as well as all individual study groups and the scientific advisory board of the Decide-TB project: Elizabeth Maleche Obimbo (chair), Stephen M. Graham, Moorine P. Sekkade, Andrew Copas, Sabine Verkuijl, Jenny Hill, Anna Scardigli, Anne K. Detjen, Albert Kuaté, Sharon Musakanya, and Charity Habeenzu.

## Contributors

OM, MB, CC, LO, JAS, PJD and MMZ conceptualised and acquired the funding for the Decide-TB study. LO, MMZ, MdE, PJD, MP, AR, RD, MHTNN, ACH, NH, HJZ, NEN, CK, MN, VPV, MB, EW, BN, RM, JM, AM, GB, JVT, LB, PG, HSS, OM, JAS, and CC contributed to original study implementation, data collection and curation. AG, LO and MMZ undertook the analysis with input from PJD, MP, MHTNN, MdE, PD, JAS, and MMZ. AG, LL, RD, MHTNN had access to and reviewed the source data. PG, JM, and HSS reviewed chest x-rays. AG, MMZ and LO wrote the first draft with input from PJD, OM, and JAS. All authors reviewed and contributed to subsequent drafts. All authors read and approved the final manuscript.

## Ethics

All original studies obtained both ethical approval as well as documented individual consent from caregivers (and assent from children, if applicable) prior to any study-specific procedures.

## Supplemental Methods

### Study Design, population, and establishment of an IPD

In the TB-Speed HIV cohort (n=72; 10.6% of the entire IPD), data regarding night sweats was not collected. A simple imputation strategy based on a decision tree model was employed. The children recruited to the TB-Speed HIV, RaPaed-TB, and Umoya studies were recruited from tertiary level facilities, whereas children recruited to the TB-Speed Decentralization study were recruited from district and primary health care centres. A dataset including only the children living with HIV (from studies which are similar in inclusion criteria to TB-Speed HIV) from RaPaed-TB (n=148), Umoya (n=42), and TB-Speed HIV (n=204) was generated; and with the simputation package in R (version 0.2.8), we used a classification decision tree approach to predict night sweats status based on clinically relevant variables (age, history of tuberculosis exposure, severe acute malnutrition, cough ≥2 weeks, fever ≥2 weeks, weight loss, cavitations on chest X-ray, and Xpert result [Mtb positive or Mtb negative]) using the function impute_cart() [1]. The imputed dataset was then used for subsequent analyses.

### Prediction score development

The prediction model was developed for children under 10 years, reflecting the target age range of the WHO treatment decision algorithms (TDAs), even though WHO eligibility criteria for the 4-month treatment regimen extend to children under 16 years.

Final models were derived using backward stepwise selection based on the Akaike Information Criterion (AIC). This corresponds to retaining predictors with likelihood-ratio test statistics more than twice the degrees of freedom, approximately equivalent to using p-value thresholds of 0.157 for 1 degree of freedom and 0.135 for 2 degrees of freedom, as recommended for clinical prediction modelling to reduce overfitting and optimism [2]. We included 11 identical categorical clinical predictors: age (3 months to 2 years; 2 years to 5 years; 5 years to 10 years), sex (female; male), Weight/BMI for Age Z-Score (>-3 to <-2; >-2 to <-1; >-1), cough lasting ≥2 weeks (Yes; No), fever lasting ≥2 weeks (Yes; No), history of tuberculosis (TB) exposure (Yes; No), HIV status (Positive; Negative), night sweats (Yes; No), palpable lymph nodes (Yes; No), tachycardia based on age (Yes; No), tachypnoea based on age (Yes; No) (Supplemental table 3). Model 1 included all 11 predictors and additionally bacteriological testing including, smear microscopy (Positive; Negative) or semiquantitative Xpert MTB/RIF or Ultra (Positive: medium, high or very high; Negative: negative, trace, very low, low) results. Model 2 included only the 11 clinical predictors.

For translation into a score, we selected a target sensitivity of >70% to balance missed severe cases against the impact of excessive false positives in low-prevalence settings. Higher sensitivity targets (e.g. ≥85%) were considered but were not selected because they lie at the extreme upper tail of the ROC curve, where small changes in predicted risk result in large shifts in specificity and classification leading to instability and a marked loss of performance. Therefore, a moderate-sensitivity operating point was selected to support practical triage decisions while maintaining robustness after translation from the regression model to the integer score.

---

**Supplemental Table 1.** Box 5.3 Eligibility criteria for the 4-month regimen (2HRZ(E)/2HR) in children and adolescents aged between 3 months and 16 years, in various settings. Adapted from the WHO operational handbook [3]. For this study, ‘WHO criteria’ refers to Setting B and Setting C.

|  | Setting | Criteria |
| --- | --- | --- |
| Eligibility criteria for the 4-month regimen (2HRZ(E)/2HR) in children and adolescents aged between 3 months and 16 years, in various settings | **A**<br>In children and adolescents who have undergone Bacteriological testing and CXR, a 4-month treatment regimen should be started in children and adolescents meeting **all of the following three criteria** | (1) CXR findings consistent with non-severe TB. CXR should ideally be done at baseline:<br>a. intrathoracic lymph node TB without significant airway obstruction;<br>OR<br>b. PTB confined to one lobe with no cavities and no miliary pattern;<br>OR<br>c. uncomplicated pleural effusion (without pneumothorax or empyema)<br>(2) Xpert MTB/RIF or Ultra is negative, trace, very low or low, (Xpert MTB/RIF <Medium) or smear-negative (if Xpert MTB/RIF or Ultra not available)<br>(3) the child has mild symptoms that do not require hospitalization. |
|  | **B**<br>In settings without CXR, children and adolescents who have undergone Bacteriological testing, a 4-month treatment regimen should be started in children and adolescents meeting **the following criteria** | (1) TB that is negative, trace, very low or low using Xpert MTB/RIF or Ultra (Xpert MTB/RIF <Medium), or smear-negative (if Xpert MTB/RIF or Ultra not available); AND<br>(2) the child or adolescent has mild symptoms that do not require hospitalization.<br>OR<br>(1) Isolated extra-thoracic (peripheral) lymph node TB, without confirmed or suspected involvement of other extrapulmonary sites of disease; AND<br>(2) the child or adolescent has mild symptoms that do not require hospitalization. |
|  | **C**<br>In settings without CXR and Bacteriological testing, a 4-month treatment regimen should be started in children and adolescents meeting **either of the following criteria** | (1) Isolated extra-thoracic (peripheral) lymph node TB, without confirmed or suspected involvement of other extrapulmonary sites of disease; AND<br>(2) the child has mild symptoms that do not require hospitalization.<br>OR<br>(1) the child has a clinical diagnosis of pulmonary TB; AND<br>(2) the child has mild symptoms that do not require hospitalization. |

Mild symptoms that do not require hospitalization include the absence of danger or high-priority signs, absence of asymmetrical or persistent wheezing, and none of the following: SAM, respiratory distress, high fever (>39°C), severe pallor, restlessness, irritability.

**Supplemental Table 2:** Study characteristics of individual studies included in the IPD [1,4].

|  | RaPaed-TB | Umoya | TB-Speed decentralization | TB-Speed HIV |
| --- | --- | --- | --- | --- |
| Aim | To evaluate several novel index tests and diagnostic approaches | To i) build a clinical, radiological, and biological sample repository to evaluate new emerging diagnostic tools and biomarkers and ii) investigate the short and long-term impact of pulmonary TB and other respiratory illnesses on lung health and QoL | To assess the impact of implementing decentralized diagnostic approaches on childhood TB case detection at district hospital (DH) and primary health centres (PHCs) and to compare decentralization strategies | To externally validate the PAANTHER score in CLHIV with presumptive TB |
| Design | Prospective single-gate diagnostic accuracy study | Prospective observational cohort study | Cross-sectional operational research study with nested prospective cohort | Prospective diagnostic cohort for external validation of TDA |
| Population | Children < 15 years with presumptive TB | Children < 13 years with presumptive TB | Children <15 years with presumptive TB | CLHIV <15 years with presumptive TB |
| Duration | Participant recruitment was conducted between January 21, 2019, and July 1, 2021 | Participant recruitment was conducted from November 2017 to May 2023 | During study intervention period (March 7, 2020 to September 30, 2021), we decentralized the comprehensive childhood TB diagnosis package and documented TB diagnosis. The exact start and end dates varied across countries. | Participant recruitment was conducted between October 2, 2019 and December 31, 2021 |

| Setting | South Africa,<br>Mozambique, Malawi,<br>India, Tanzania | South Africa | Cambodia, Cameroon,<br>Côte d'Ivoire,<br>Mozambique, Sierra<br>Leone, Uganda | Côte d'Ivoire,<br>Mozambique, Uganda,<br>Zambia |
| --- | --- | --- | --- | --- |
| Number screened | 5,313 | 1,330 | 584 | 713 |
| Number recruited | 975 | Up to 600 children with<br>presumptive pulmonary TB<br>and 100 healthy controls | 3,104 / 584 | 277 |
| Inclusion criteria | Consent and assent (if<br>applicable), confirmation<br>of TB disease and/or signs<br>and symptoms suggestive<br>of TB | Consent and assent (if<br>applicable), <13 years,<br>> 2.5 kg, presumptive<br>pulmonary TB | For assessing<br>decentralized diagnostic:<br>Sick children seeking care<br>at OPD of DH or PHC, age<br><15 years<br>For comparing<br>decentralization<br>strategies: Age <15 years,<br>Presumptive TB defined as<br>children presenting ≥1<br>systematic screening<br>criterion<sup>1</sup>, Presumptive TB<br>identified by the site<br>clinician irrespective of the<br>above criteria, especially<br>presumed extra-<br>pulmonary TB cases,<br>Informed consent signed,<br>Child's assent obtained in<br>those aged >7 years | Children aged 1 month to<br>14 years, Documented<br>HIV-infection, Presumptive<br>TB based on at least one<br>criteria<sup>4</sup>, History of<br>contact with a TB case and<br>any of the symptoms<sup>2</sup> with<br>a shorter duration (< 2<br>weeks), Informed consent<br>signed by parent/guardian |
| Exclusion criteria | Critical condition posing<br>an undue risk to the child,<br>body weight < 2 Kg, age ≥<br>15 years, currently<br>receiving anti-TB drugs | Extra-thoracic TB only, TB<br>treatment for >2 days in<br>previous 2 weeks,<br>Alternative diagnosis at<br>baseline, Severe illness<br>resulting in unstable<br>clinical condition, Contra- | For assessing<br>decentralized diagnostic:<br>None<br>For comparing<br>decentralization<br>strategies: Children who | Ongoing TB treatment or<br>history of intake of anti-TB<br>drugs in the last 3 months<br>(isoniazid alone or<br>rifampin/isoniazid for<br>preventive therapy is not<br>an exclusion criteria) |
|  |  | indication to sampling procedures, Discharge before baseline sampling completed, Unstable social circumstances, Residence in remote areas | have received TB treatment in the past 6 months |  |
| Chest X-ray review for reference variable | CXR's were reviewed by single expert | CXR's were reviewed by single expert | CXR's were reviewed by single expert | CXR's were reviewed by single expert |

<sup>1</sup> Cough with a duration of >2 weeks, Fever with a duration of >2 weeks, Documented weight loss, History of TB contact with any duration of cough
<sup>2</sup> Persistent cough for more than 2 weeks, persistent fever for more than 2 weeks, recent failure to thrive (documented clear deviation from a previous growth trajectory in the last 3 months or Z score weight/age < 2), failure of broad-spectrum antibiotics for treatment of pneumonia, suggestive CXR features
Abbreviations: TB: Tuberculosis, QoL: Quality of Life, OPD: outpatient department, PHC: primary health care level, DH: district health level, CLHIV; Children Living with HIV, TDA: Treatment Decision Algorithm.

**Supplemental Table 3:** Study Variables [1].

| Variable | Definitions | RaPaed | UMOYA | TB-Speed HIV | TB-Speed Decentralisation |
| --- | --- | --- | --- | --- | --- |
| Age groups | 3 months to <2 years; 2 years to <5 years; 5 years to 10 years | Date of birth or age (years) | Date of birth or age (years) | Date of birth or age (years) | Date of birth or age (years) |
| Cough ≥ 2 weeks | Persistent, unremitting cough for 2 weeks or more | Presence of cough at enrolment (binary) and cough duration | Presence of cough at enrolment and classification into acute (incl. number of days) or chronic (>2 weeks) | Presence of cough in the 4 weeks preceding enrolment (binary) and duration | Presence of cough > 2 weeks at enrolment (binary) |
| Weight/BMI for Age (Z score) | Anthropometric Z-scores for weight-for-age and BMI-for-age calculated from measured weight and height using WHO approved methods including R packages: Anthro for children <5 years; Anthroplus for children ≥5 years | Weight-for-age Z-score for children <5 years and BMI-for-age Z-score for children aged 5–10 years (continuous) | Weight-for-age Z-score for children <5 years and BMI-for-age Z-score for children aged 5–10 years (continuous) | Weight-for-age Z-score for children <5 years and BMI-for-age Z-score for children aged 5–10 years (continuous) | Weight-for-age Z-score for children <5 years and BMI-for-age Z-score for children aged 5–10 years (continuous) |
| Fever ≥ 2 weeks | Persistent fever for 2 weeks or more (the score in the algorithm is based on the duration of fever as per the history rather than the actual temperature on examination) | Presence of fever at enrolment (binary) and duration of fever | Presence of fever at enrolment (binary) and duration of fever | Presence of fever in the 4 weeks preceding enrolment (binary) and duration | Presence of fever > 2 weeks at enrolment (binary) |
| Night sweats | Excessive night-time sweating that soaks the bed or clothes | Presence of night sweats at enrolment (binary) | Presence of night sweats at enrolment (binary) | **Missing/not collected** | Presence of night sweats at enrolment (binary) only in children $\geq 5$ years |
| Swollen lymph nodes | Non-painful, enlarged cervical, submandibular or axillary lymph nodes | Presence of lymph node at enrolment (binary) which are not characterised as painful, are bigger than 1cm, and are cervical, axillar, or submandibular | Presence of lymphadenopathy at enrolment (binary) which are bigger than 1.5cm, and are cervical, axillar, or submandibular | Presence of cervical or supra-clavicular Adenopathy (no, single, multiple) | Presence of adenopathy at enrolment (binary) with specified location (supra-clavicular or cervical), number, and size of biggest node (1-3 cm or >3 cm) |
| Tachycardia | Children aged under 2 months: heart rate over 160 beats/minute<br>Children aged 2–12 months: heart rate over 150 beats/minute<br>Children aged 12 months to 5 years: heart rate over 140 beats/minute | Children aged under 2 months: heart rate over 160 beats/minute<br>Children aged 2–12 months: heart rate over 150 beats/minute<br>Children aged 12 months to 5 years: | Children aged under 2 months: heart rate over 160 beats/minute<br>Children aged 2–12 months: heart rate over 150 beats/minute<br>Children aged 12 months to 5 years: | Children aged under 1 year: heart rate over 160 beats/minute<br>Children aged 1–2 years: heart rate over 150 beats/minute<br>Children aged 2-5 years: heart rate over 140 beats/minute | Children aged under 2 months: heart rate over 160 beats/minute<br>Children aged 2–12 months: heart rate over 150 beats/minute<br>Children aged 12 months to 5 years: |
|  | Children aged over 5 years:<br>heart rate over 120<br>beats/minute | heart rate over 140<br>beats/minute<br>Children aged over 5<br>years: heart rate over<br>120 beats/minute | heart rate over 140<br>beats/minute<br>Children aged over 5<br>years: heart rate over<br>120 beats/minute | Children aged over 5<br>years: heart rate over<br>120 beats/minute | heart rate over 140<br>beats/minute<br>Children aged over 5<br>years: heart rate over<br>120 beats/minute |
| Tachypnoea | Children aged under 2<br>months: respiratory rate<br>over 60/minute<br>Children aged 2–12<br>months: respiratory rate<br>over 50/minute<br>Children aged 12 months to<br>5 years: respiratory rate<br>over 40/minute<br>Children aged over 5 years:<br>respiratory rate over<br>30/minute | Children aged under 2<br>months: respiratory<br>rate over 60/minute<br>Children aged 2–12<br>months: respiratory<br>rate over 50/minute<br>Children aged 12<br>months to 5 years:<br>respiratory rate over<br>40/minute<br>Children aged over 5<br>years: respiratory rate<br>over 30/minute | Children aged under 2<br>months: respiratory<br>rate over 60/minute<br>Children aged 2–12<br>months: respiratory<br>rate over 50/minute<br>Children aged 12<br>months to 5 years:<br>respiratory rate over<br>40/minute<br>Children aged over 5<br>years: respiratory rate<br>over 30/minute | Not collected but<br>respiratory rate and<br>age available | Children aged under 2<br>months: respiratory<br>rate over 60/minute<br>Children aged 2–12<br>months: respiratory<br>rate over 50/minute<br>Children aged 12<br>months to 5 years:<br>respiratory rate over<br>40/minute<br>Children aged over 5<br>years: respiratory rate<br>over 30/minute |
| HIV status |  | Overall HIV status<br>generated based on<br>reported status and<br>test results. Overall<br>HIV test results based<br>on the combination of<br>the first test with the second and third confirmatory test | Overall HIV status<br>generated based on<br>reported status and<br>test results | Entire cohort is living<br>with HIV based on<br>inclusion criteria for<br>the study | HIV status following<br>testing and in<br>children < 18 months,<br>an additional PCR<br>was done |
| SAM |  | Based on weight for length Z-score and BMI-for-age Z-score < -3 (weight for length used in those under 5) | If weight for length < -3 Z-score or bilateral pitting oedema for those under 5 months of age<br>If weight for length < -3 Z-score or MUAC < 11.5 cm (with or without oedema for those over 5 months) | BMI-for-age Z-score < -3 (weight for height in those under 5); MUAC; presence of oedema | Weight for height Z-score < -3 if child aged < 5 years or binary indication of severe acute malnutrition |
| Collected respiratory specimen |  | Sample type indicated as spontaneous or induced sputum, nasopharyngeal aspirate, gastric aspirate, or other respiratory sample | Indication if two respiratory samples were collected and if not, how many were collected | Sample type indicated as expectorated sputum, gastric aspirate or nasopharyngeal aspirate | Indication of expectorated sputum collection and of nasopharyngeal aspirate collection (binary) |
| Xpert conducted |  | Type of Xpert test (MTB-Rif or MTB Rif Ultra) | Type of Xpert test (MTB-Rif or MTB Rif Ultra) | Type of Xpert test (MTB-Rif or MTB Rif Ultra) | Type of Xpert test (MTB-Rif or MTB Rif Ultra) |
| Mtb final result |  | Final Xpert result (positive or negative). This is the highest result as multiple samples were collected per child | Mtb complex detected or trace result | Mtb complex detected or trace result | Mtb complex detected or trace result |
| TB exposure < 12 months |  | TB exposure at 24 months | Indication of TB exposure in the past 12 months | Indication of TB exposure in the past 12 months | Indication of TB exposure in the past year |
| **Variables for CXR evaluation** |  |  |  |  |  |
| Cavities |  | Presence of cavities at enrolment evaluated by central expert reviewer (binary) | Presence of cavities at enrolment evaluated by central expert reviewer (binary) | Presence of cavities at enrolment evaluated by central expert reviewer (binary) | Presence of cavities at enrolment evaluated by by central expert reviewer (binary) |
| Enlarged lymph nodes |  | Presence of enlarged lymph nodes at enrolment evaluated by central expert reviewer (binary) | Presence of enlarged lymph nodes at enrolment evaluated by central expert reviewer (binary) | Presence of cavities at enrolment evaluated by central expert reviewer (binary) | Presence of enlarged lymph nodes at enrolment evaluated by central expert reviewer (binary) |
| Opacities |  | Presence of opacities at enrolment evaluated by central expert reviewer (binary) | Presence of opacities at enrolment evaluated by central expert reviewer (binary) | Presence of interstitial opacities at enrolment evaluated by central expert reviewer (binary) | Presence of opacities at enrolment evaluated by central expert reviewer (binary) |
| Miliary pattern |  | Presence of miliary patterns at enrolment | Presence of miliary patterns at enrolment | Presence of miliary at enrolment evaluated | Presence of miliary patterns at enrolment |
|  |  | evaluated by central expert reviewer (binary) | evaluated by central expert reviewer (binary) | by central expert reviewer (binary) | evaluated by central expert reviewer (binary) |
| Effusion |  | Presence of effusion at enrolment evaluated by central expert reviewer (binary) | Presence of effusion at enrolment evaluated by central expert reviewer (binary) | Presence of pleural, pericardial or peritoneal effusion at enrolment evaluated by central expert reviewer (binary) | Presence of effusion at enrolment evaluated by central expert reviewer (binary) |
| Airway compression |  | **Missing/not collected** | **Missing/not collected** | Presence of large airway compression at enrolment evaluated by central expert reviewer (binary) | Presence of large airway compression at enrolment evaluated by central expert reviewer (binary) |
| Diagnostic classification |  | Based on NIH criteria (2015) | Based on NIH criteria (2015) | Based on NIH criteria (2015) | Based on NIH criteria (2015) |

Most variables were able to be used or minorly manipulated to fit the WHO definitions to be used in the TDA. There were two cases where changes had to be made; 1) as TB contact was recorded in variations including no time specification, within 12 months, or within 24 months, for this study we took any TB contact as positive. 2) Night sweats was entirely missing/not collected in the TB-Speed HIV cohort and we thus conducted simple imputation based on a logistic regression to address the missingness [1].
Abbreviations: SAM: severe acute malnutrition, NIH: National Institutes of Health, Mtb: mycobacterium tuberculosis, BMI: Body Mass Index, PCR: polymerase chain reaction, MUAC: mid upper arm circumference, WFA: weight-for-age, TDA: Treatment Decision Algorithm

**Supplemental Table 4.** Outcome definitions used in study, based on NIH case definitions [5].

|  | Categories | Definitions |
| --- | --- | --- |
| TB classification | Unlikely | Bacteriological confirmation NOT obtained AND criteria for “unconfirmed tuberculosis” NOT met OR children that met “unconfirmed tuberculosis” but did not receive anti-tuberculosis treatment and were well at follow-up |
|  | Unconfirmed | Bacteriological confirmation NOT obtained AND at least 2 of the following:<br>• Symptoms/signs suggestive of tuberculosis<sup>1</sup> (as defined)<br>• Chest radiograph<sup>2</sup> consistent with tuberculosis<br>• Close tuberculosis exposure or immunologic evidence of *Mtb* infection<br>• Positive response to tuberculosis treatment (requires documented positive clinical response on tuberculosis treatment - no time duration specified) |
|  | Confirmed | Bacteriological confirmation obtained - requires *Mtb* to be confirmed (culture, Xpert MTB/RIF, or Xpert MTB/Rif Ultra assay) from at least 1 respiratory specimen |

<sup>1</sup>Symptoms/signs suggestive of TB include cough for more than 2 weeks, fever for more than 2 weeks, and poor weight gain or weight loss in the past 3 months. In young children (< 5 years), reduced playfulness or lethargy is also included.
<sup>2</sup>Chest X-rays were assessed by study-specific clinicians or external experts and defined as “attributable to TB”, “unlikely TB”, or “normal”. According to the operational handbook, common abnormalities in children include enlarged hilar and paratracheal lymph nodes, alveolar consolidation without visible cavities, miliary lesions, and pleural effusions [3].
Abbreviations: *Mtb*, *mycobacterium tuberculosis*. NIH, National Institute of Health.

**Supplemental Table 5.** Score 1 development for Model 1, with bacteriological testing.

| Predictors | Level | OR | Model $\beta$ coefficient | Final Predictor Score (x scaling factor: 38.2) |
| --- | --- | --- | --- | --- |
| History of TB exposure | Yes | 0.7 | -0.3 | -13 |
| HIV status positive | Yes | 1.6 | 0.5 | 18 |
| Night sweats | Yes | 0.6 | -0.5 | -20 |
| Tachycardia | Yes | 1.6 | 0.5 | 19 |
| Tachypnoea | Yes | 1.9 | 0.7 | 26 |
| Bacteriological Testing | Positive<br>(Xpert MTB/RIF ≥Medium or smear-positive) | 7.1 | 1.9 | 75 |
| Intercept |  | - | -1.9 | -75.8 |
| Predicted probability cutoff, $P$ | | - | 0.15 | 5.8 |
| Logit, $P$ | | - | -1.723 | -65.8 |
| Severity diagnosis threshold<br>(Logit [$P$] – intercept) | | - | 0.36 | 10.0 |

OR, odds ratio; $P$ : Predicted probability cutoff; Xpert MTB/RIF ≥ medium – medium, high, very high.

**Supplemental Table 6.** Score 2 development for Model 2, without bacteriological testing.

| Predictors | Levels | OR | Model $\beta$<br>coefficient | Final Predictor<br>Score (× scaling<br>factor: 79.4) |
| --- | --- | --- | --- | --- |
| History of TB exposure | Yes | 1.2 | -0.4 | -29 |
| Night sweats | Yes | 0.7 | -0.4 | -28 |
| Tachycardia | Yes | 1.7 | 0.5 | 43 |
| Tachypnoea | Yes | 2.0 | 0.7 | 57 |
| Intercept |  | - | -1.69 | -134.5 |
| Predicted probability<br>cutoff, $P$ | | - | 0.17 | 13.7 |
| Logit, $P$ | | - | -1.568 | -124.5 |
| Severity diagnosis<br>threshold<br>(Logit [$P$] – intercept) | | - | 0.13 | 10.0 |

OR, odds ratio; $P$ : Predicted probability cutoff;

**Supplemental Table 7.**
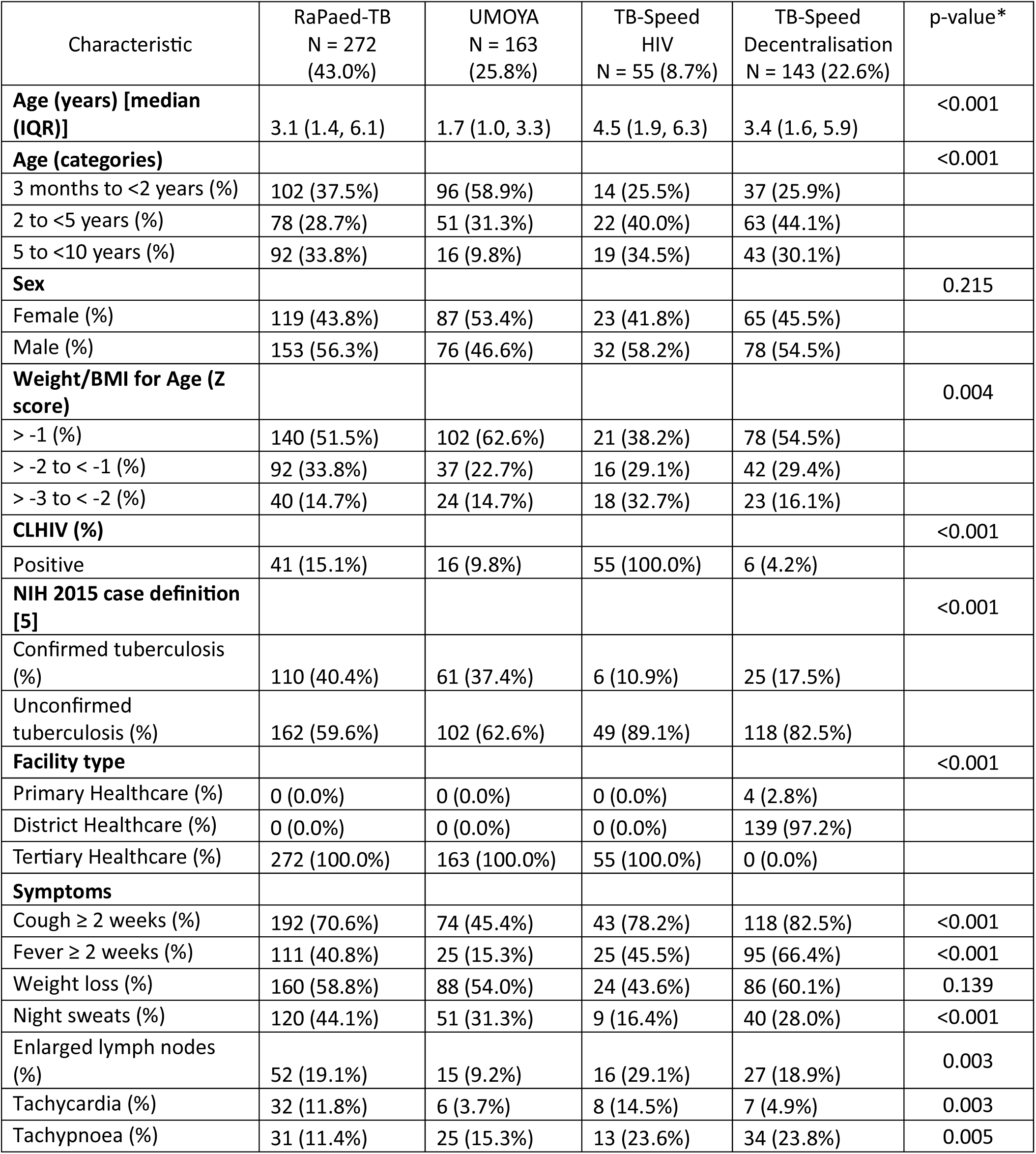
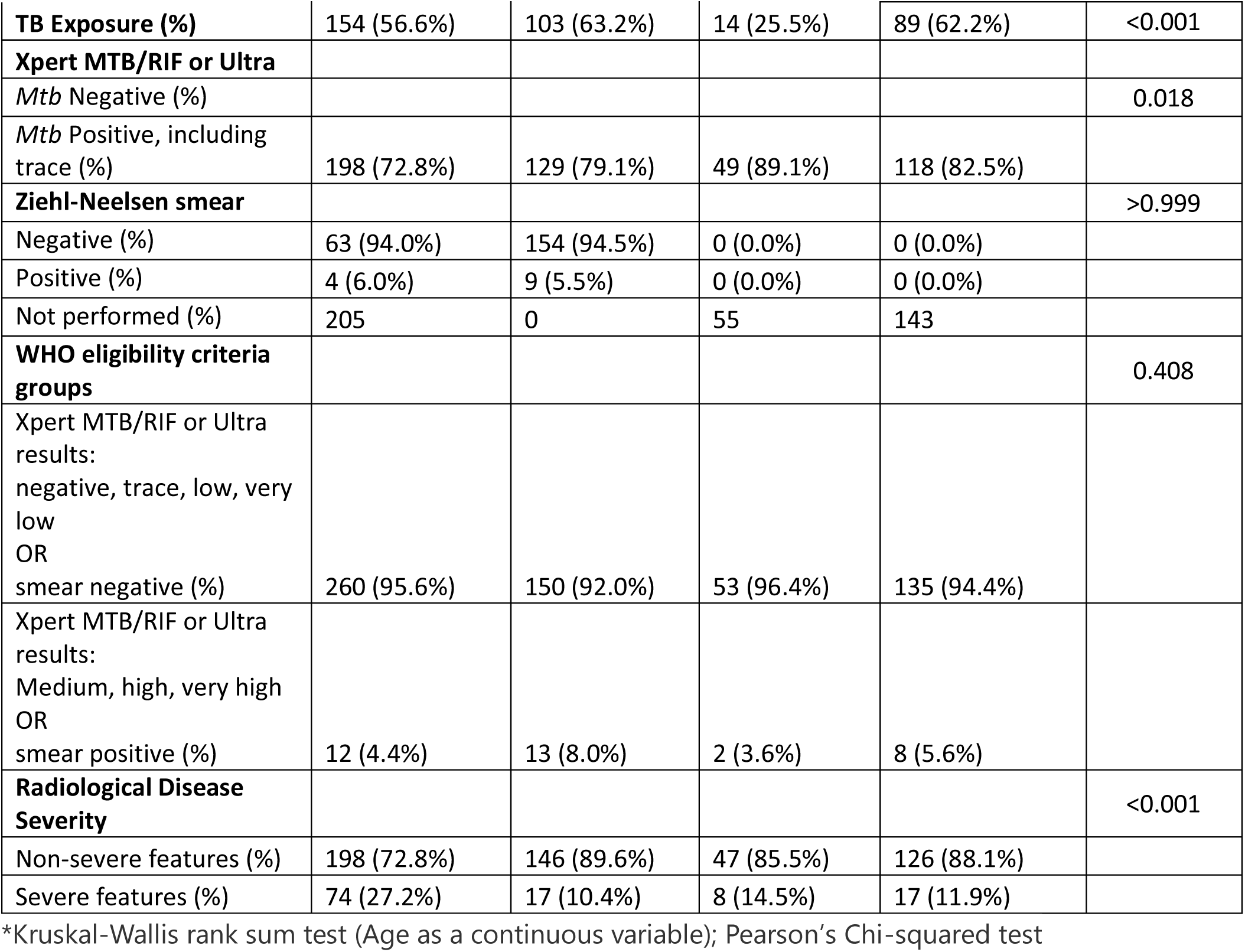
Characteristics of children included in the individual patient dataset by parent study.

**Supplemental Table 8.** Diagnostic accuracy of the WHO-criteria where bacteriological testing available and in subgroups against radiological severity as the reference standard.

|  | TP | FP | FN | TN | Sensitivity (95% CI)* |  | Specificity (95% CI)* |  |
| --- | --- | --- | --- | --- | --- | --- | --- | --- |
| Where CXR is unavailable and bacteriological testing is available |  |  |  |  |  |  |  |  |
| WHO-criteria | 25 | 57 | 91 | 460 | 30.1% | 20.3 – 40.2 | 83.4% | 80.2 – 86.4 |
| Study |  |  |  |  |  |  |  |  |
| RaPaed-TB | 11 | 18 | 63 | 180 | 14.9% | 7.0–23.6 | 90.9% | 86.6–94.8 |
| UMOYA | 6 | 12 | 11 | 134 | 35.4% | 12.5–60.9 | 91.7% | 86.7–95.9 |
| TB-Speed Dec. | 6 | 19 | 11 | 107 | 34.5% | 12.5–57.1 | 85.0% | 78.5–91.0 |
| TB-Speed HIV | 2 | 8 | 6 | 39 | 24.5% | 0.0–60.0 | 83.0% | 71.2–93.6 |
| NIH 2015 diagnosis |  |  |  |  |  |  |  |  |
| Confirmed TB | 19 | 25 | 34 | 119 | 36% | 22.9–50 | 82.60% | 76.1–88.8 |
| Unconfirmed TB | 6 | 27 | 57 | 322 | 9.50% | 2.9–17.5 | 92.30% | 89.2–95.1 |
| Age |  |  |  |  |  |  |  |  |
| 3 months to <2 years | 13 | 28 | 38 | 170 | 25.5% | 13.2–37.5 | 85.8% | 80.7–90.9 |
| 2 to <5 years | 4 | 14 | 32 | 164 | 10.9% | 2.4–21.7 | 92.2% | 88.1–95.9 |
| 5 to <10 years | 8 | 15 | 21 | 126 | 27.2% | 10.7–44.0 | 89.3% | 84.0–94.5 |
| Weight for Age Z Score |  |  |  |  |  |  |  |  |
| > -1 | 11 | 19 | 47 | 264 | 19.0% | 9.1–29.3 | 93.2% | 90.1–95.9 |
| > -2 to < -1 | 5 | 16 | 29 | 137 | 14.4% | 3.2–26.9 | 89.4% | 84.4–94.0 |
| > -3 to < -2 | 9 | 22 | 15 | 59 | 37.0% | 17.9–56.3 | 72.9% | 63.1–82.1 |
| HIV status |  |  |  |  |  |  |  |  |
| Children living with HIV | 5 | 11 | 22 | 80 | 18.4% | 4.2–34.4 | 88.0% | 80.4–94.4 |
| Children without HIV | 20 | 46 | 69 | 380 | 22.3% | 14.0–31.8 | 89.2% | 86.2–92.0 |
| Healthcare Level |  |  |  |  |  |  |  |  |
| District Healthcare | 6 | 17 | 11 | 105 | 34.4% | 11.8–57.2 | 86.1% | 79.7–92.0 |
| Tertiary Healthcare | 19 | 38 | 80 | 353 | 19.4% | 12.0–27.5 | 90.3% | 87.4–93.1 |
| TB contact History |  |  |  |  |  |  |  |  |
| History of TB exposure | 8 | 33 | 46 | 273 | 14.8% | 6.1–24.6 | 89.2% | 85.7–92.5 |
| No history of TB exposure | 17 | 24 | 45 | 187 | 27.3% | 16.7–39.0 | 88.6% | 84.2–92.5 |

\*Mean sensitivity and specificity after bootstrapping 1000 times.
TB: Tuberculosis; TP: True Positives; TN: True Negatives; FP: False Positives; FN: False Negatives. NIH 2015 diagnosis: National Institute of Health (NIH 2015) clinical case definitions; CI: Confidence Intervals; WHO: World Health Organisation.

**Supplemental Table 9.**
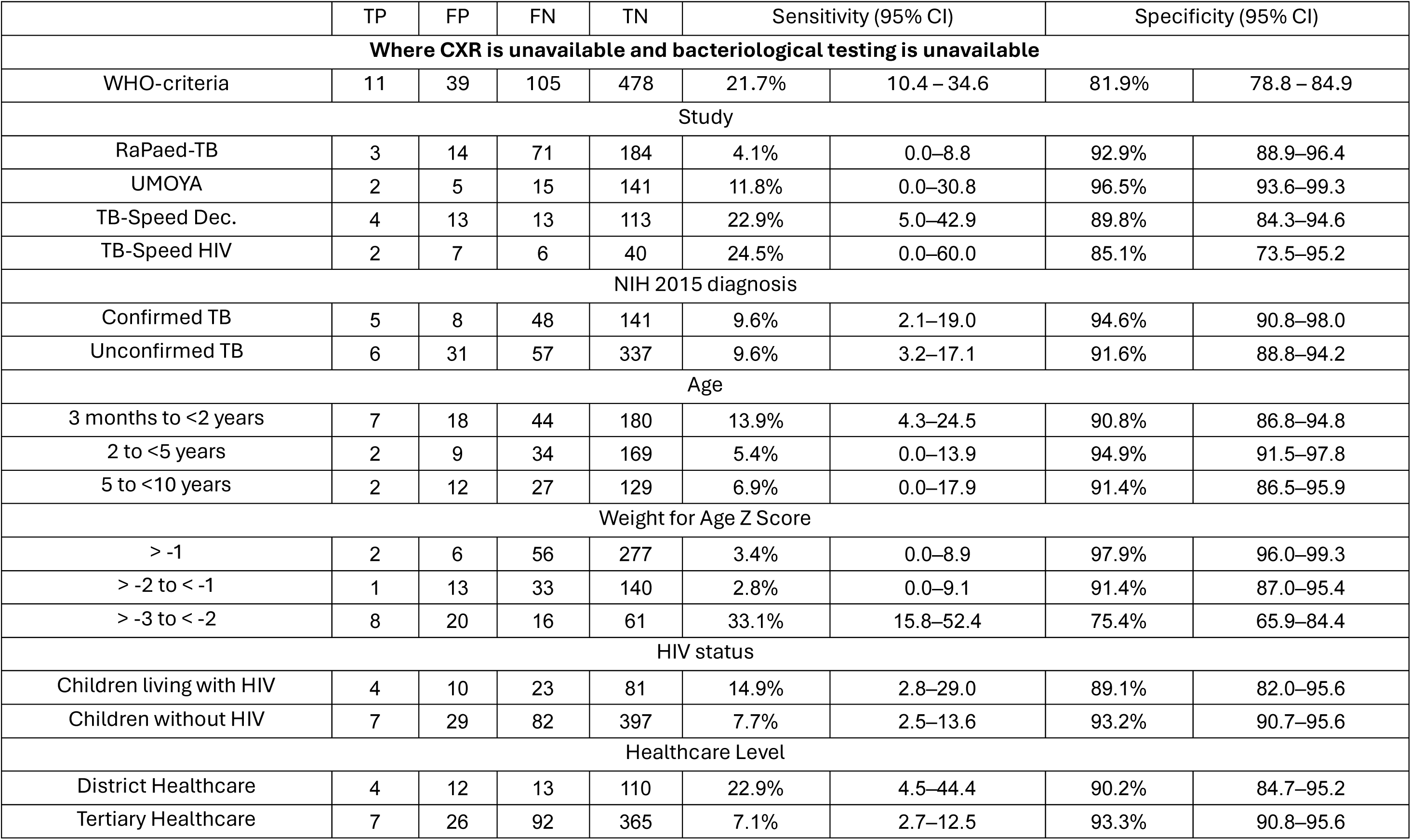
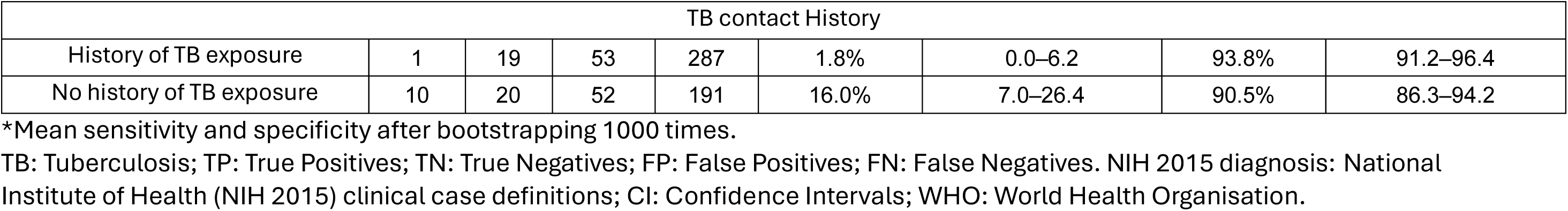
Diagnostic accuracy of the WHO-criteria where bacteriological testing unavailable and in subgroups against radiological severity as the reference standard.

**Supplemental Figure 1.**
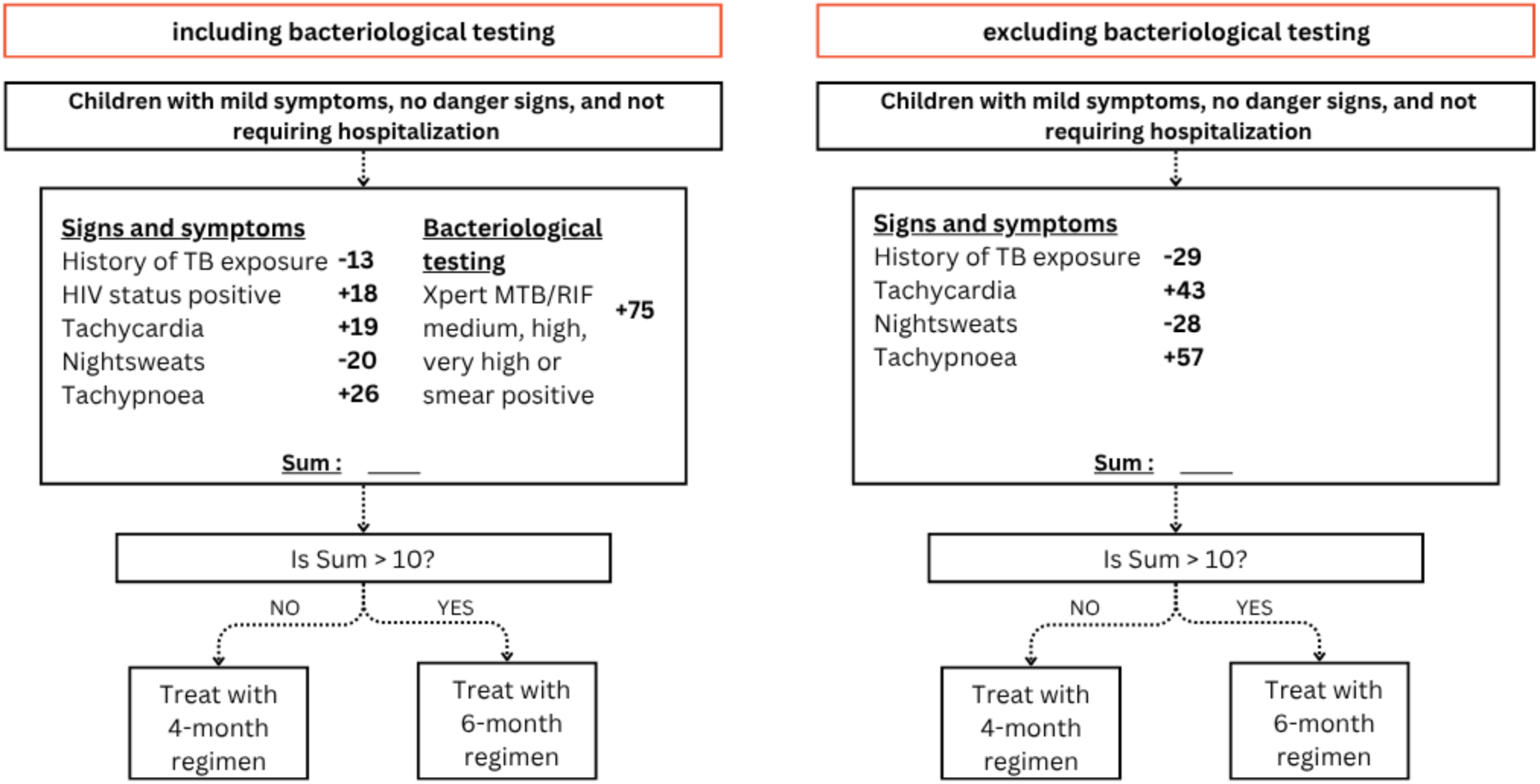
Scoring system schematic for possible use in practice for different settings alongside the WHO Treatment Decision Algorithms in the absence of chest X-ray. Here score 1 is based on model 1 and refers to absence of chest X-ray, but with bacteriological testing. Score 2 is based on model 2 and refers to absence of chest X-ray and without bacteriological testing.

**Supplemental Table 10.** Diagnostic accuracy of the developed score with bacteriological testing for predicting CXR severity in different subgroups.

|  | TP | FP | FN | TN | Sensitivity (95% CI)* |  | Specificity (95% CI)* |  |
| --- | --- | --- | --- | --- | --- | --- | --- | --- |
| Where CXR is unavailable and bacteriological testing is available |  |  |  |  |  |  |  |  |
| Score 1 | 48 | 117 | 68 | 400 | 41.1% | 32.4-49.5 | 77.3% | 73.6-80.8 |
| Study |  |  |  |  |  |  |  |  |
| RaPaed-TB | 24 | 30 | 50 | 168 | 32.5% | 22.2–43.1 | 84.9% | 80.0–89.7 |
| UMOYA | 10 | 29 | 7 | 117 | 59.3% | 33.3–83.3 | 80.1% | 73.5–86.4 |
| TB-Speed Dec. | 9 | 25 | 8 | 101 | 53.1% | 31.2–77.8 | 80.0% | 73.5–86.7 |
| TB-Speed HIV | 5 | 33 | 3 | 14 | 63.0% | 25.0–100.0 | 30.1% | 17.9–44.4 |
| NIH 2015 diagnosis |  |  |  |  |  |  |  |  |
| Confirmed TB | 25 | 36 | 28 | 113 | 47.1% | 33.0–59.6 | 75.9% | 68.6–82.7 |
| Unconfirmed TB | 23 | 81 | 40 | 287 | 36.5% | 25.0–47.9 | 78.2% | 74.0–82.2 |
| Age |  |  |  |  |  |  |  |  |
| 3 months to <2 years | 23 | 49 | 28 | 149 | 45.7% | 31.8–59.6 | 75.2% | 69.3–80.6 |
| 2 to <5 years | 11 | 36 | 25 | 142 | 30.2% | 15.4–45.0 | 79.7% | 73.3–85.6 |
| 5 to <10 years | 14 | 32 | 15 | 109 | 47.3% | 28.8–66.2 | 77.2% | 71.0–83.7 |
| Weight for Age Z Score |  |  |  |  |  |  |  |  |
| > -1 | 24 | 50 | 34 | 233 | 41.8% | 28.9–54.5 | 82.2% | 77.9–86.6 |
| > -2 to < -1 | 12 | 39 | 22 | 114 | 35.1% | 20.0–51.7 | 74.2% | 67.3–80.8 |
| > -3 to < -2 | 12 | 28 | 12 | 53 | 50.6% | 28.8–70.8 | 65.2% | 54.7–74.7 |
| HIV status |  |  |  |  |  |  |  |  |
| Children living with HIV | 15 | 56 | 12 | 35 | 56.0% | 37.7–73.2 | 38.5% | 28.5–48.8 |
| Children without HIV | 33 | 61 | 56 | 365 | 37.3% | 27.8–47.5 | 85.7% | 82.2–88.9 |
| Healthcare Level |  |  |  |  |  |  |  |  |
| District Healthcare | 9 | 23 | 8 | 99 | 53.4% | 29.4–76.9 | 81.0% | 74.2–87.7 |
| Tertiary Healthcare | 39 | 92 | 60 | 299 | 39.4% | 29.5–49.3 | 76.6% | 71.9–80.7 |
| TB contact History |  |  |  |  |  |  |  |  |
| History of TB exposure | 15 | 36 | 39 | 270 | 28.0% | 16.8–40.2 | 88.1% | 84.4–91.7 |
| No history of TB exposure | 33 | 81 | 29 | 130 | 53.3% | 41.6–66.1 | 61.7% | 55.9–68.6 |

\*Mean sensitivity and specificity after bootstrapping 1000 times.
TB: Tuberculosis; TP: True Positives; TN: True Negatives; FP: False Positives; FN: False Negatives. NIH 2015 diagnosis: National Institute of Health (NIH 2015) clinical case definitions; CI: Confidence Intervals.

**Supplemental Table 11.** Diagnostic accuracy of the developed scores without bacteriological testing for predicting CXR severity in different subgroups.

|  | TP | FP | FN | TN | Sensitivity (95% CI)* |  | Specificity (95% CI)* |  |
| --- | --- | --- | --- | --- | --- | --- | --- | --- |
| Where CXR is unavailable and bacteriological testing is unavailable |  |  |  |  |  |  |  |  |
| Score 2 | 36 | 88 | 80 | 429 | 30.9% | 22.6–40.4 | 83.0% | 79.5–86.3 |
| Study |  |  |  |  |  |  |  |  |
| RaPaed-TB | 19 | 29 | 55 | 169 | 25.9% | 15.8–36.9 | 85.3% | 80.4–90.0 |
| UMOYA | 5 | 21 | 12 | 125 | 30.0% | 8.3–53.6 | 85.7% | 79.7–91.2 |
| TB-Speed Dec. | 8 | 24 | 9 | 102 | 47.3% | 25.0–71.8 | 80.8% | 73.5–87.7 |
| TB-Speed HIV | 4 | 14 | 4 | 33 | 49.9% | 15.4–85.2 | 70.4% | 56.0–82.7 |
| NIH 2015 diagnosis |  |  |  |  |  |  |  |  |
| Confirmed TB | 14 | 19 | 39 | 130 | 26.3% | 14.7–38.2 | 87.4% | 82.1–92.3 |
| Unconfirmed TB | 22 | 69 | 41 | 299 | 34.8% | 24.0–47.5 | 81.4% | 77.6–85.0 |
| Age |  |  |  |  |  |  |  |  |
| 3 months to <2 years | 18 | 39 | 33 | 159 | 35.4% | 21.8–50.0 | 80.2% | 74.3–85.6 |
| 2 to <5 years | 10 | 23 | 26 | 155 | 27.3% | 13.1–42.2 | 86.9% | 81.1–91.6 |
| 5 to <10 years | 8 | 26 | 21 | 115 | 27.3% | 12.0–45.3 | 81.8% | 75.4–88.1 |
| Weight for Age Z Score |  |  |  |  |  |  |  |  |
| > -1 | 15 | 37 | 43 | 246 | 25.8% | 15.7–38.1 | 86.9% | 82.9–90.7 |
| > -2 to < -1 | 11 | 29 | 23 | 124 | 31.7% | 16.4–48.3 | 80.7% | 74.5–86.5 |
| > -3 to < -2 | 10 | 22 | 14 | 59 | 42.3% | 22.6–61.9 | 72.7% | 63.0–81.4 |
| HIV status |  |  |  |  |  |  |  |  |
| Children living with HIV | 9 | 23 | 18 | 68 | 32.7% | 15.8–50.0 | 74.7% | 65.3–83.1 |
| Children without HIV | 27 | 65 | 62 | 361 | 30.5% | 21.5–39.8 | 84.7% | 81.5–87.9 |
| Healthcare Level |  |  |  |  |  |  |  |  |
| District Healthcare | 8 | 23 | 9 | 99 | 47.4% | 23.5–72.5 | 81.0% | 74.0–87.7 |
| Tertiary Healthcare | 28 | 64 | 71 | 327 | 28.3% | 20.0–38.3 | 83.7% | 79.8–87.4 |

| TB contact History |  |  |  |  |  |  |  |  |
| --- | --- | --- | --- | --- | --- | --- | --- | --- |
| History of TB exposure | 11 | 29 | 43 | 277 | 20.5% | 10.3–32.5 | 90.4% | 87.1–93.6 |
| No history of TB exposure | 25 | 59 | 37 | 152 | 40.6% | 28.6–54.2 | 72.2% | 65.4–77.6 |

\*Mean sensitivity and specificity after bootstrapping 1000 times.
TB: Tuberculosis; TP: True Positives; TN: True Negatives; FP: False Positives; FN: False Negatives. NIH 2015 diagnosis: National Institute of Health (NIH 2015) clinical case definitions; CI: Confidence Intervals.

